# The methodological quality of COVID-19 systematic reviews is low, except for Cochrane reviews: a meta-epidemiological study

**DOI:** 10.1101/2020.08.28.20184077

**Authors:** Yuki Kataoka, Shiho Oide, Takashi Ariie, Yasushi Tsujimoto, Toshi A. Furukawa

## Abstract

**Objectives:** The objective of this study was to investigate the methodological quality of COVID-19 systematic reviews (SRs) indexed in medRxiv and PubMed, compared with Cochrane COVID Reviews.

**Study Design and Setting:** This is a cross-sectional meta-epidemiological study. We searched medRxiv, PubMed, and Cochrane Database of Systematic Reviews for SRs of COVID-19. We evaluated the methodological quality using A MeaSurement Tool to Assess systematic Reviews (AMSTAR) checklists. The maximum AMSTAR score is 11, and minimum is 0. Higher score means better quality.

**Results:** We included 9 Cochrane reviews as well as randomly selected 100 non-Cochrane reviews in medRxiv and PubMed. Compared with Cochrane reviews (mean 9.33, standard deviation 1.32), the mean AMSTAR scores of the articles in medRxiv were lower (mean difference -2.85, 95%confidence intervals (CI): -0.96 to -4.74) and those in PubMed was also lower (mean difference -3.28, 95% CI: -1.40 to -5.15), with no difference between the latter two.

**Conclusions:** It should be noted that AMSTAR is not a perfect tool of assessing quality SRs other than intervention. Readers should pay attention to the potentially low methodological quality of COVID-19 SRs in both PubMed and medRxiv but less so in Cochrane COVID reviews.

**PROTOCOL AND REGISTRATION:** We developed the protocol before conducting this study (Kataoka Y, Oide S, Ariie T, Tsujimoto Y, Furukawa TA. Quality of COVID-19 research in preprints: a meta-epidemiological study protocol. Protocols.io 2020. https://doi.org/10.17504/protocols.io.bhm8j49w.).

**What is new?:**

Key findings
- The methodological quality of COVID-19 systematic reviews (SRs) in medRxiv and PubMed were lower than Cochrane COVID reviews.
- The methodological quality of reviews in medRxiv and PubMed did not differ.

What this study adds to what was known
- Expert opinions and a preliminary review suggested the low quality of COVID-19 SRs but this hypothesis has not been examined empirically.
- We evaluated the methodological quality of COVID-19 SRs using comprehensive search and confirmed that the quality was low except for Cochrane reviews.

**What is the implication and what should change now?:** Readers should pay attention to the potentially low methodological quality of COVID-19 SRs in both PubMed and medRxiv but less so in Cochrane COVID reviews.

The methodological quality of COVID-19 SRs except for Cochrane COVID reviews needed to be improved.

## 1. Background

SARS-Cov-2 virus has caused a once-in-a-century pandemic. As of August 2020, there have been over 800,000 coronavirus disease 2019 (COVID-19) deaths [1]. Recent advances in information and communication technology have led to the publication of many academic articles [2,3]. Until May 2020, more than a thousand COVID-19 trials were registered in ClinicalTrials.gov [4]. Cochrane started Cochrane COVID Reviews to answer the time-sensitive needs of health decision makers as fast as possible. These reviews are intended to simultaneously assure the scientific quality [5].

While the number of studies is growing, questions are being raised about their methodological quality. A meta-epidemiological study that investigated the quality of randomized controlled studies (RCT) of COVID-19 pointed out the poor methodology [6]. A preliminary meta-epidemiological study including 18 COVID-19 systematic reviews (SRs) published until March 2020 also pointed out the poor methodology [7]. Despite the important role of SRs in clinical decision-making [8], the quality of COVID-19 SRs performed for speedy reporting has not been adequately evaluated. In addition, no studies assessed the quality of SRs published in preprint servers without peer review in comparison with other data sources. Hence, we investigated the methodological quality of COVID-19 reviews indexed in medRxiv, PubMed, and the Cochrane Library.

## 2. Materials and methods

### 2.1 protocol and study design

This is a cross-sectional meta-epidemiological study. We abode by the reporting guideline of meta-epidemiological study where applicable (S1 Table) [9]. We published the protocol prior to conducting this study [10].

### 2.2 Types of studies included

We included SRs, indexed in PubMed, medRxiv, and Cochrane Database of Systematic Reviews (CDSR). We included articles of topics related to the COVID-19 practice, irrespective of publication types. We included Cochrane Reviews that dealt with the COVID-19 pandemic. We excluded study protocols.

The definition of SRs was “a scientific investigation that focuses on a specific question and uses explicit, prespecified scientific methods to identify, select, assess, and summarize the findings of similar but separate studies.” [11]

### 2.3 Search methods

We retrieved abstracts from medRxiv COVID-19 SARS-CoV-2 preprints using search terms: “review”, “evidence synthesis”, “meta-analysis”, or “metaanalysis” on 15^th^ June 2020. [12] We retrieved abstracts from PubMed using Shokraneh’s filter for COVID-19 [13] and PubMed Systematic Reviews Filter [14] on 15^th^ June 2020. We retrieved abstracts from Cochrane Database of Systematic Reviews (CDSR) using search term “COVID-19” on 17^th^ Aug 2020.

### 2.4 Study selection

Two of three review authors (YK, SO, and TA) selected abstracts from search results independently. Disagreements were solved through discussion. Two of three review authors (YK, SO, and TA) selected full text articles from selected abstracts independently. Disagreements were solved through discussion. Of the articles indexed in medRxiv and PubMed and meeting the eligibility criteria, we randomly selected 100 from each database for inclusion in the present study. We included all Cochrane reviews.

### 2.5 Data extraction and assessment

#### 2.5.1 Methodological quality of systematic reviews

We defined the methodological quality as “to what extent a study was designed, conducted, analyzed, interpreted, and reported to avoid systematic errors” [15].

For calibration training, three review authors (YK, SO, and TA) independently evaluated the methodological quality using A MeaSurement Tool to Assess systematic Reviews (AMSTAR) checklists for five included articles [16]. Disagreements were resolved through discussion. Then one of three review authors (YK, SO, and TA) evaluated other articles. Another author (YK or SO) confirmed the results. We resolved disagreements through discussion.

We initially intended to use AMSTR 2, which is a critical appraisal tool for systematic reviews that include randomized or non-randomized studies of healthcare interventions [17]. Because there were few intervention reviews, we used AMSTAR due to the latter’s applicability [14]. AMSTAR was developed for assessing the methodological quality of systematic reviews. For each of the 11 items in AMSTAR checklist, we calculated the AMSTAR score by counting the number of “Yes”. Higher scores indicate higher quality of the systematic review. The possible maximum score was 11. We added some explanations to AMSTAR to reduce disagreements following a previous study after calibration training [18]. The details are shown in S2 Table.

#### 2.5.2 Other variables

The number of included studies was counted at the time of their synthesis of each article. We evaluated types of research questions, the presence of protocol registrations, and the presence of limitation in each abstract. We determined that articles with “rapid” in the title is a “rapid review”. We also evaluated the presence of SPIN in the title or abstract conclusion in intervention systematic reviews whose first outcome was non-significant. [19] SPIN was judged present when there were the manipulation of language to potentially mislead readers from the likely truth of the results. One of three review authors (YK, SO, and TA) evaluated these variables. Another author (YK or SO) confirmed the results. We resolved the disagreements through discussion.

#### 2.5.3 Outcomes

Our primary outcome was the total score of AMSTAR.

### 2.6 Data analysis

We described summary statistics. We used risk difference (RD) and 95% confidence intervals (CI) to compare binary variables. For quantification of the disagreements of AMSTAR checklists, we calculated kappa statistics between the initial and the final evaluations for the included studies except for the five articles used for calibration. We used ANOVA with Bonferroni correction for the comparison of total AMSTAR score. We conducted sensitivity analysis excluding scoping reviews or qualitative synthesis, because the summary score will be lower in those not intended for meta-analysis or risk-of-bias assessment. We used Stata ver. 16.1 (StataCorp LLC, College Station, Texas, United States of America).

### 2.7 Ethics

Because this study used only open data, ethics approval was not applicable.

## 3. Results

### 3.1 Results of the search

The details of selection process are shown in Figure 1. We searched a total of 641 abstracts from medRxiv and PubMed. We randomly included 49 articles from medRxiv, and 51 articles from PubMed, respectively. We searched a total of 12 abstracts from CDSR, and included 9 articles. Detailed citations are shown in S3 Table. Included articles from PubMed was not indexed in medRxiv, or CDSR.

**Fig 1.**
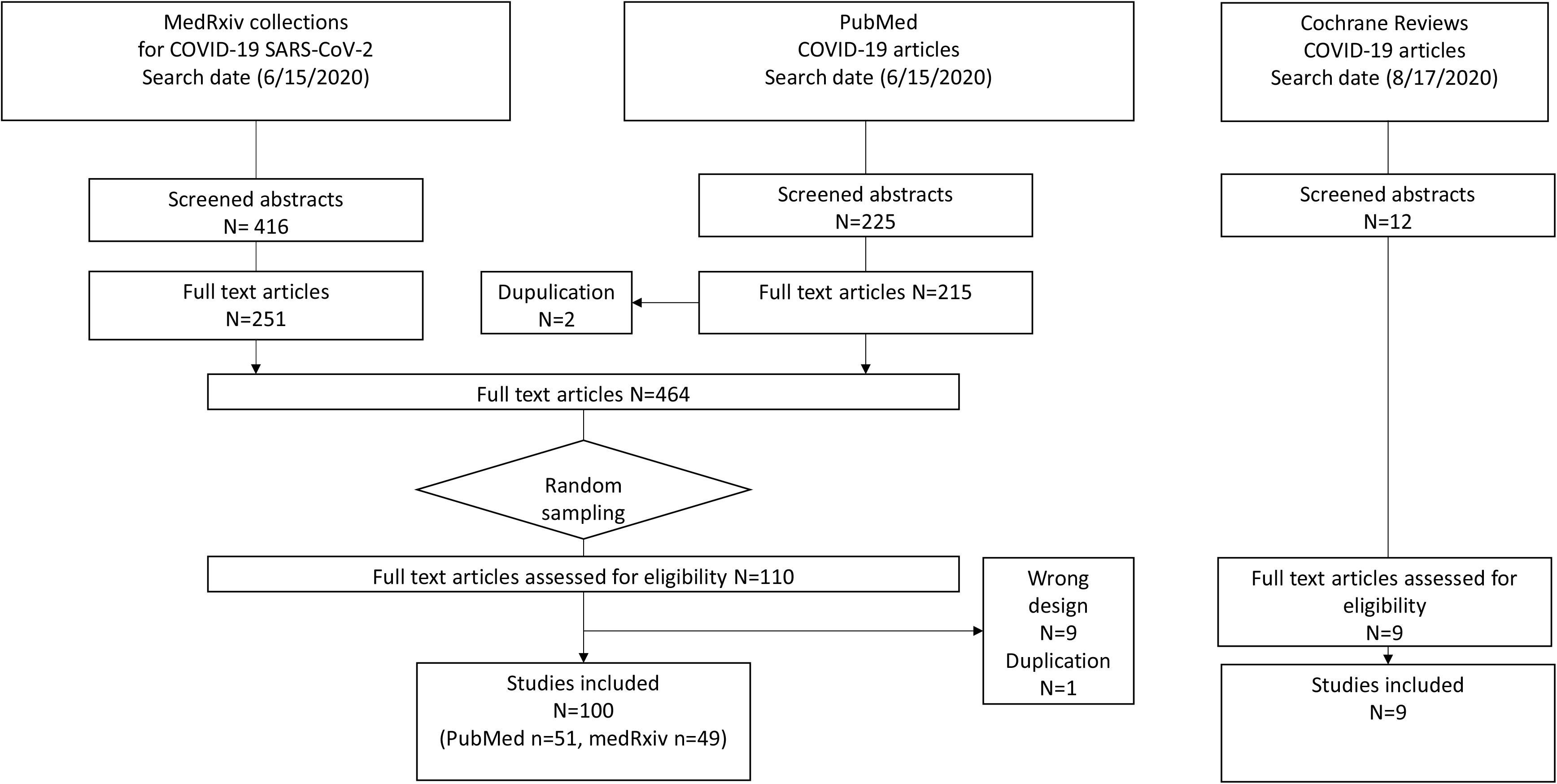
Flowchart for the selection

### 3.2 Characteristics of included articles

The characteristics of included articles are shown in Table 1. We included 7 rapid reviews from medRxiv, 4 from PubMed, and 4 from Cochrane, respectively. The medians [interquartile range (IQR)] of included studies for qualitative synthesis were 29.5 [17-45.5] in medRxiv, 18.5 [11-39] in PubMed, and 24 [16-36] in Cochrane, respectively. More than a half of articles did not register their protocols.

**Table 1.** characteristics of included systematic reviews

|  | medRxiv<br>N=49 | PubMed<br>N=51 | Cochrane<br>N=9 |
| --- | --- | --- | --- |
| Number of included articles | 29.5 (17-45.5) | 18.5 (11-39) | 24 (16-36) |
| Median (IQR) |  |  |  |
| Types of research questions |  |  |  |
| DTA | 1 (2%) | 2 (4%) | 2 (22%) |
| Intervention | 9 (18%) | 14 (28%) | 6 (67%) |
| Meta-epidemiological | 1 (2%) | 0 (0%) |  |
| Prevalence, incidence | 8 (16%) | 16 (31%) |  |
| Prognostic factor | 11 (22%) | 7 (14%) |  |
| Scoping review, qualitative synthesis | 19 (39%) | 12 (24%) | 1 (11%) |
| Protocol registration |  |  |  |
| Present | 5 (10%) | 9 (18%) | 3 (33%) |

IQR: interquartile range, DTA: diagnostic test accuracy

### 3.3 Quality of included articles

Table 2 summarizes the characteristics of included articles. The agreement between the initial and the agreed-upon AMSTAR ratings ranged between 0.85 and 1.0. The mean (standard deviation (SD)) scores was 9.33 (1.32) in Cochrane reviews, 6.48 (2.07) of medRxiv, and 6.06 (2.30) of PubMed, respectively. Referring to limitations in each abstract were more often in Cochrane than medRxiv (RD 56%, 95% CI: 32 to 81), and also more often in Cochrane than PubMed (RD 64%, 95%CI: 41 to 88), respectively. Compared with Cochrane reviews, the mean scores of the reviews in medRxiv were lower (mean difference -2.85, 95%confidence intervals (CI): -0.96 to -4.74) and those in PubMed was also lower (mean difference -3.28, 95% CI: -1.40 to -5.15) (Table 3). The difference between medRxiv and PubMed was not statistically significant. The score difference between articles in medRxiv and those in PubMed were not statistically significant (mean difference -0.42, 95% CI: -1.46 to 0.62). Sensitivity analysis after excluding scoping reviews or qualitative synthesis showed similar difference (Cochrane vs. medRxiv: mean difference -2.71, 95%CI 0.70 to 4.72, Cochrane vs. PubMed: mean difference -2.93, 95%CI 0.98 to 4.87).

**Table 2.** The quality of included systematic review articles

|  | medRxiv | PubMed | Cochrane | Risk difference<br>medRxiv vs.<br>Cochrane<br>% [95% CI] | Risk difference<br>PubMed vs.<br>Cochrane | kappa |
| --- | --- | --- | --- | --- | --- | --- |
| Referring to limitations in each abstract |  |  |  |  |  | 1 |
| Yes | 16 (33%) | 12 (24%) | 8 (89%) | 56% [32 to 81] | 64% [41 to 80] |  |
| No | 33 (67%) | 37 (73%) | 1 (11%) |  |  |  |
| Without an abstract | 0 (0%) | 2 (4%) | 0 (0%) |  |  |  |
| AMSTAR |  |  |  |  |  |  |
| 1. Was an 'a priori' design provided? |  |  |  |  |  | 0.91 |
| Yes | 8 (17%) | 13 (25%) | 8 (89%) | 72% [49 to 95] | 63% [40 to 88] |  |
| No | 4 (8%) | 1 (2%) | 0 (0%) |  |  |  |
| Can't answer | 36 (75%) | 38 (73%) | 1 (11%) |  |  |  |
| 2. Was there duplicate study selection and data extraction? |  |  |  |  |  | 0.96 |
| Yes | 41 (85%) | 45 (87%) | 9 (100%) | 15% [5 to 25] | 13% [4 to 23] |  |
| No | 1 (2%) | 3 (6%) | 0 (0%) |  |  |  |
| Can't answer | 6 (13%) | 4 (8%) | 0 (0%) |  |  |  |
| 3. Was a comprehensive literature search performed? |  |  |  |  |  | 1 |
| Yes | 46 (96%) | 47 (90%) | 7 (78%) | -18% [-46 to 10] | -13% [-41 to 16] |  |
| No | 2 (4%) | 5 (10%) | 2 (22%) |  |  |  |
| 4. Was the status of publication (i.e. grey literature) used as an inclusion criterion? |  |  |  |  |  | 0.85 |
| Yes | 21 (44%) | 24 (46%) | 6 (67%) | 23% [-11 to 57] | 21% [-13 to 54] |  |
| No | 18 (38%) | 14 (27%) | 3 (33%) |  |  |  |
| Can't answer | 8 (17%) | 14 (27%) | 0 (0%) |  |  |  |
| Not applicable | 1 (2%) | 0 (0%) | 0 (0%) |  |  |  |
| 5. Was a list of studies (included and excluded) provided? |  |  |  |  |  | 0.96 |
| Yes | 5 (10%) | 2 (4%) | 8 (89%) | 78% [56 to 100] | 85% [64 to 100] |  |
| No | 43 (90%) | 50 (96%) | 1 (11%) |  |  |  |
| 6. Were the characteristics of the included studies provided? |  |  |  |  |  | 0.85 |
| Yes | 47 (98%) | 47 (90%) | 9 (100%) | 2% [-2 to 6] | 10% [2 to 18] |  |
| No | 1 (2%) | 5 (10%) | 0 (0%) |  |  |  |
| 7. Was the scientific quality of the included studies assessed and documented? |  |  |  |  |  | 0.96 |
| Yes | 27 (56%) | 22 (42%) | 9 (100%) | 44% [30 to 58] | 58% [44 to 71] |  |
| No | 21 (44%) | 30 (58%) | 0 (0%) |  |  |  |
| 8. Was the scientific quality of the included studies used appropriately in formulating conclusions? |  |  |  |  |  | 0.98 |
| Yes | 31 (65%) | 24 (46%) | 9 (100%) | 35% [22 to 49] | 54% [40 to 67] |  |
| No | 17 (35%) | 28 (54%) | 0 (0%) |  |  |  |
| 9. Were the methods used to combine the findings of studies appropriate? |  |  |  |  |  | 0.89 |
| Yes | 24 (50%) | 22 (42%) | 3 (33%) | -17% [-51 to 17] | -9 [-43 to 25] |  |
| No | 4 (8%) | 2 (4%) | 1 (11%) |  |  |  |
| Not applicable | 20 (42%) | 28 (54%) | 5 (56%) |  |  |  |
| 10. Was the likelihood of publication bias assessed? |  |  |  |  |  | 0.96 |
| Yes | 22 (46%) | 21 (40%) | 7 (78%) | 32% [1 to 63] | 37% [7 to 68] |  |
| No | 25 (52%) | 31 (60%) | 2 (22%) |  |  |  |
| Not applicable | 1 (2%) | 0 (0%) | 0 (0%) |  |  |  |
| 11 . Was the conflict of interest stated? |  |  |  |  |  | 0.96 |
| Yes | 39 (81%) | 48 (92%) | 9 (100%) | 19% [8 to 30] | 8% [0 to 15] |  |
| No | 9 (19%) | 4 (8%) | 0 (0%) |  |  |  |

95%CI: 95% confidence intervals
We calculated kappa statistics for the initial evaluation and final evaluation.

**Table 3.** The difference of AMSTAR score

|  | Mean (SD) | Mean difference [95%CI] |
| --- | --- | --- |
| Cochrane | 9.33 (1.32) | Reference |
| medRxiv | 6.48 (2.07) | -2.85 [-0.96 to -4.74] |
| PubMed | 6.06 (2.30) | -3.28 [-1.40 to -5.15] |

We used ANOVA with Bonferroni correction.

There were six intervention articles whose first presented outcome was not statistically significant. No articles expressed SPIN in both titles and conclusions of abstracts.

## 4. Discussion

This is the first meta-epidemiological study evaluating methodological quality of SRs dealing with the COVID-19 pandemic including preprints, peer-reviewed articles, and Cochrane reviews. Compared with Cochrane reviews, the mean scores of AMSTAR were lower in articles from medRxiv and PubMed, respectively. The mean scores of medRxiv and PubMed did not differ significantly.

Our findings suggest ordinary peer reviews could not improve the quality of SRs. The mean AMSTAR score difference between medRxiv (6.06) and PubMed (6.48) were not statistically significant. A previous study included SRs published in PubMed from China and US showed their mean AMSTAR score was 6.14 [20]. There’s not much difference between our results and regular peer-reviewed SRs. This fact suggests it’s not clear whether the quality of COVID-19 SRs will improve in the future with sufficient time of peer-review. At this point, readers should note that the methodological quality of SRs about COVID-19 in both PubMed and medRxiv may be of poor quality, but this is not the case in the Cochrane COVID reviews.

Strict structured reporting like Cochrane reviews would be more useful than time-limited peer reviews or without peer reviews [21]. Referring to limitations in each abstract were more often in Cochrane than in medRxiv or PubMed. The differences of the quality of the articles we found between Cochrane reviews and others included the presentation of protocols, listing included and excluded studies, considering the quality of included studies at the conclusions, examining publication bias, and presenting the conflict of interests. These domains will be improved by the appropriate use of preferred reporting items for systematic reviews and meta-analyses (PRISMA) statement [22,23]. If the duplicate study selection is difficult due to manpower insufficiency, one solution might be to use crowdsourcing to bring people together through the internet [24].

The proportion of protocol registrations did not differ from those reported in previous studies. Previous studies reported about 20% of SRs registered their protocols [25,26]. Protocol registration is important to prevent duplicate efforts, prevent outcome reporting bias, and to reduce alpha errors in the results of meta-analyses. PROSPERO, the largest SRs protocol registration site, has some problems including not accepting Scoping reviews and taking more than 30 days to register [27]. Some SRs published their protocols in preprint servers [28,29]. For speedy and assuring the scientific quality, protocol registration in preprint servers would be useful.

Our study has several limitations. First, AMSTAR is a reliable and valid measurement tool that has been used widely so far [30]. However, it was originally developed for SRs of randomized trials [31], and there is a criticism for the integrated evaluation of different domains [32]. Future research is needed to determine how to assess the quality of SRs which do not target at interventions. Second, we assessed the quality of the articles while not being masked about the published journal names. While the empirical evidence shows that such a bias is unlikely [33], this information bias could have led to overestimating the quality of Cochrane COVID reviews. Third, we were not able to incorporate studies that were published before and after peer review in the current study. Further study to compare studies published in peer-reviewed journals after publication in preprint servers is warranted.

## 5. Conclusions

Readers should pay attention to the potentially low methodological quality of SRs about COVID-19 in both PubMed and medRxiv but less so in Cochrane COVID reviews. The methodological quality of COVID-19 SRs except for Cochrane COVID reviews needed to be improved.

## Data Availability

Data will be shared on inquiries.

## Author Contributions

Yuki Kataoka had full access to all of the data in the study and take responsibility for the integrity of the data and the accuracy of the data analysis.

Concept and design: All authors.

Acquisition, analysis, or interpretation of data: All authors.

Drafting of the manuscript: Yuki Kataoka.

Critical revision of the manuscript for important intellectual content: All authors.

Supervision: Toshi A. Furukawa.

## Additional Contributions

We acknowledge Ms. Kyoko Wasai, who assisted retrieving full text articles.

## SUPPORTING INFORMATION CAPTIONS

S1 Table: Items for reporting methodological research, adapted from the PRISMA Checklist

S2 Table Modified AMSTAR checklist

S3 Table retrieved full text articles and decision

